# Treatment and Cost -outcomes of a simplified antiretroviral treatment strategy for hepatitis C among HCV and HIV co-infected patients in Ukraine

**DOI:** 10.1101/2021.03.19.21253780

**Authors:** Sergie Antoniak, C Charles Chasela, Morgan J Freiman, Yulia Stopolianska, Tetiana Barnard, MM Gandhi, Maria Liulchuk, Zhanna Tsenilova, V Tretiakov, Jeri Dible, Constance Wose Kinge, T Minior, Sofiane Mohamed, Fadzai Marange, Clint Cavenaugh, Charles van der Horst, Svitlana Antonyak, Thembisile Xulu, Kara W Chew, Ian Sanne, Sydney Rosen, for EQUIP Health

## Abstract

**Background:** We conducted a demonstration project of an integrated HIV and viral hepatitis testing and treatment strategy using generic ledipasvir/sofosbuvir (LDV/SOF).

**Methods:** Eligible HCV viraemic adults from two clinics in Kyiv were treated with LDV/SOF +/-weight-based ribavirin for 12 weeks. Clinical assessments were performed at screening and week 24 and as needed; treatment was dispensed every 4 weeks. The primary outcome was sustained virologic response (SVR)12 weeks after treatment. Program costs in 2018 USD were estimated per patient treated using observed resource utilization, local unit, and antiretroviral therapy (ART) costs over the 24-week period.

**Results:** 868 participants initiated on treatment, 87% (755) were PWID and 55.5% (482) were HIV co-infected. The common genotypes were 1 (74.1%) and 3 (22%) and SVR was achieved in 831/868 (95.7%) by intention-to-treat analysis. The average cost per patient treated was $680, assuming generic LDV/SOF and ribavirin pricing and standard quantitative HCV viral load testing. Medications comprised 38% of the average cost/patient, laboratory tests 26%, events (clinic visits, counselling) 10%, and indirect costs 26%. ART accounted for 60% of all drug costs, with HCV medications just 40%.

**Conclusion:** Generic LDV/SOF +/- ribavirin provided produced exceptionally good outcomes including amongst patients with genotype 3 HCV and PWID at an average cost of <$700/patient year, including ART for those with HIV. Under the assumptions of generic drug pricing but higher laboratory costs, an average cost of $750/patient is likely a reasonable estimate for this intervention in Ukraine, excluding costs for scaling or maintaining the treatment program.

## Introduction

Globally, an estimated 71 million people are living with chronic hepatitis C virus (HCV) infection and are at risk for significant morbidity, including liver cirrhosis, liver failure, hepatocellular carcinoma and death^1-3^. Even in low-HCV prevalence settings, HCV infection remains common among people who inject drugs (PWID) and HIV-infected men who have sex with men (MSM)^1,4^. Despite the development of highly effective direct-acting antiviral (DAA) treatment, which can cure HCV infection with 8-12 weeks of therapy, HCV remains a leading cause of mortality worldwide, causing more than 350,000 deaths each year^5,65^. Of these, 85,000 are in Eastern Europe^7^, including Ukraine. The WHO’s Global Health Sector Strategy sets goals of a 90% reduction in new HCV infections and a 65% reduction in HCV-related mortality by 2030^8,9^. Progress in HCV treatment scale-up has been encouraging, with more than 3 million treated globally with direct-acting antivirals since 2015. Testing and diagnosis rates are still less than 10% in LMICs, however^10^.

In Ukraine, roughly 3-5% of the population—as many as 2 million people—are estimated to be HCV-infected^11^. Those most at risk are PWID, people living with HIV (PLHIV), sex workers (SW), MSM, prisoners, members of the military, populations in conflict zones, and sexual partners of PLHIV. A lack of adequate facilities and free-to-client services, combined with fear of prosecution for drug use and stigma suffered by key populations, particularly PWID and MSM,, means that most individuals who present for treatment for HCV and HIV are already well advanced in their illness^12^.

Low cost, scalable, integrated HIV and HCV testing and treatment strategies offer a chance to achieve WHO targets for HIV and HCV elimination in dual epidemic regions. We describe the treatment outcomes and estimated costs for an integrated HIV and HCV treatment program that offered HCV treatment with 12 weeks of sofosbuvir/ledipasvir (LDV/SOF) with or without weight-based ribavirin (WBR) in Ukraine.

## Patients and Methods

### Study sites and population

Enrolment into the HCV/HIV demonstration project was conducted in 2018 at two clinical facilities in Kyiv, Ukraine: the Clinic of the Gromashevsky Institute of Epidemiology and Infectious Diseases and Treatment, and Kyiv City Clinical Hospital #5. Patients were recruited to the study from HCV treatment waiting lists available at the sites, were referred by partner organizations, or had a positive HCV antibody or RNA test at one of the sites during the enrolment period. Target populations for study enrolment were PWID, MSM, SW, and sexual partners of HCV-infected individuals, though these characteristics were not required for enrolment.

### Study eligibility

Eligible participants included in the treatment study were HCV viraemic, HCV treatment naïve or experienced (prior pegylated interferon [PegIFN] and ribavirin [RBV] only), and 18 years or older, with HCV genotype 1, 2, 3, 4, 5 or 6 and with or without HIV-1 co-infection. Patients with compensated cirrhosis (Child-Pugh Class A) and hepatitis B virus (HBV) infection were eligible; those with decompensated cirrhosis (Child-Pugh Class B or C) or prior treatment with HCV DAAs were not and referred to other treatment centres for care. All participants provided written informed consent.

### Intervention

The intervention combined HCV and HIV testing, HCV treatment, simplified HCV treatment monitoring, and HIV treatment initiation for those with HIV co-infection not yet on antiretroviral therapy. HCV treatment was with fixed-dose combination ledipasvir/sofosbuvir/ (LDV/SOF) 400 mg/90 mg +/-weight-based ribavirin (1000 mg for patients <75 kg and 1200 mg for those ≥75 kg administered orally in two divided doses) for 12 weeks and provided by the project. Ribavirin was included in the regimen for treatment experienced cirrhotic genotype 1 and 4 participants and all participants with genotype 3 HCV. HIV treatment was per national guidelines, within the National HIV treatment program^13^. Participants with HBV co-infection and hepatitis B surface antigen positivity were concurrently treated with tenofovir. Participants were followed for 24 weeks (through 12 weeks after treatment completion), including assessments of HCV and HIV treatment outcomes and safety. Intervention steps are illustrated in Figure 1. A four-week supply of study medications was dispensed at weeks 0, 4, and 8 and psychosocial and peer support were provided at entry, 4, 8, 12 and 24 weeks. Participants not already on medication-assisted treatment (MAT) were referred for harm reduction, including MAT programs.

**Figure 1.**
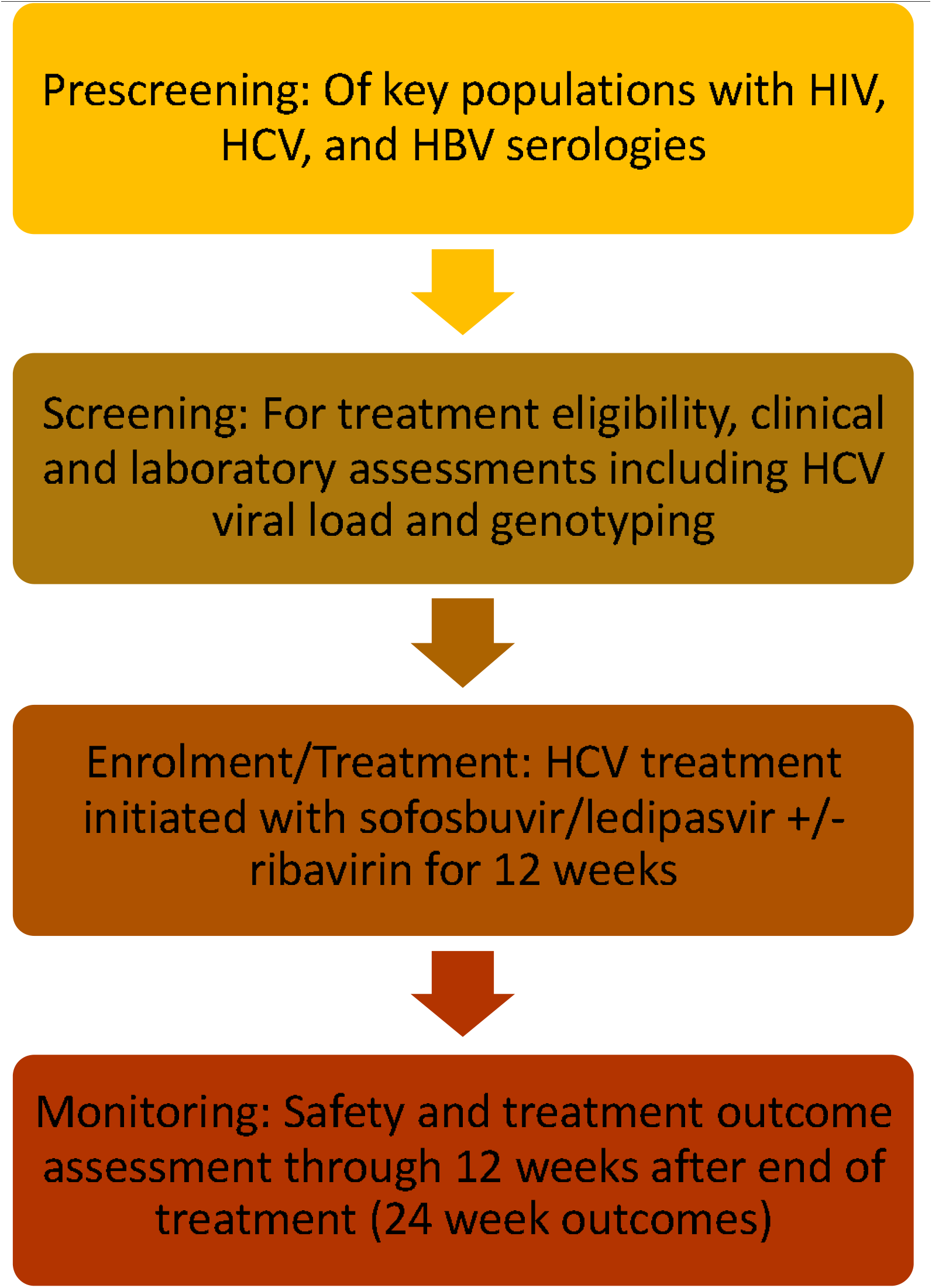
Intervention procedures. HIV = human immunodeficiency virus; HCV = hepatitis C virus; HBV = hepatitis B virus

### Clinical and laboratory evaluations

#### Clinical evaluations and drug dispensing

A complete clinical assessment including medical history, physical exam, and review of concomitant medications was performed at screening. Subsequently, clinical assessments were driven by reported symptoms and required only at entry and week 24. Liver disease stage (presence or absence of cirrhosis) was defined by any of the following and by the study physician’s judgement: cirrhosis on liver ultrasound; liver histology (liver biopsy); AST-to-platelet ratio index (APRI)>1.0 and FIB-4>3.25; or liver stiffness by transient elastography (Fibroscan) ≥ 12.5 kPa. Child-Pugh scores were calculated for all cirrhotic patients, with compensated cirrhosis defined as Child-Pugh score ≤6 (Class A).

#### Laboratory evaluations

Pre-treatment laboratory tests included HCV antibody if not already available, HCV RNA for HCV antibody positive participants, haemoglobin, platelets, aspartate, aminotransferase (AST), alanine aminotransferase (ALT), total bilirubin, albumin, creatinine, blood urea nitrogen, pregnancy testing, prothrombin time/International Normalized Ratio (INR), HBV serologies (hepatitis B core Ab (HbcAb), hepatitis B surface antigen (HbsAg), hepatitis B surface antibody (HBsAb)), and CD4+ T cell count for HIV-infected participants. Laboratory results were also obtained from the medical record if available within the specified window.

The protocol called for all participants who initiated HCV treatment and remained in care to have the following laboratory tests at week 24: HCV RNA, haemoglobin, platelets, AST, ALT, bilirubin, albumin, creatinine, blood urea nitrogen, prothrombin time/INR, and HIV RNA. Participants receiving ribavirin had haemoglobin monitoring at weeks 2, 4, and 8 and pregnancy testing (for females of childbearing potential) at weeks 4, 8, 12, and 24. Participants with eGFR<60 at screening and also receiving tenofovir disoproxil fumarate (TDF) had creatinine monitoring at weeks 4 and 12. Participants with isolated hepatitis B core Ab (HBcAb+/HBsAg-/HBsAb-) additionally had hepatic function tests (AST, ALT, total bilirubin) measured at weeks 4, 8, and 12 to monitor for HBV reactivation. Quantitative HCV viral load (real time RT-PCR reagent kit D-0794, Vector Best) and HCV genotyping (hybridization-fluorescence, AmpliSense HCV-genotype-FL reagent kit, Central Epidemiology Research Institute, Russian Federation) were performed at the Synevo central laboratory in Kyiv for all participants.

### Outcomes

The two primary outcomes of the analysis were virologic status at week 24 and cost per patient, stratified by outcome. We assigned each patient one of four virologic status outcomes at 24 weeks after treatment initiation: 1) treatment success, defined as sustained virologic response (SVR) at 12 weeks after the end of treatment (week 24); 2) treatment failure (HCV viremia greater than the lower limit of detection 12 weeks after therapy completion); 3) loss to follow up (did not return to clinic for 24-week evaluation); or 4) death (died within the 24-week study period). Other outcomes included safety (adverse events) during the treatment period.

### Data analysis

#### Outcomes

Baseline sociodemographic, clinical, and laboratory characteristics of the participants were calculated using proportions or means (standard deviations) or medians (interquartile ranges) for continuous data. The proportion with SVR was calculated for each clinical site and overall, by intention to treat (ITT). Differences between those successfully treated SVR achieved compared to those who failed treatment were assessed by t test or chi-square test as appropriate, and by multivariable logistic regression. A p-level of less than 0.2 in the bivariate analysis was used to select variables for multivariable analysis. A final model was determined using stepwise backward elimination method and only p-level of less than 0.05 were considered for the final model. Multi collinearity test and Hosmer-Lemeshow test (HL test) for logistic regression were undertaken on the final model. Data were analysed using STATA version 15 SE (StataCorp 2017. Stata Statistical Software: Release 15. College Station, TX: StataCorp LLC).

#### Costs

Costs were estimated from the provider perspective from initial screening date until assessment of cure 12 weeks after HCV treatment completion (24 weeks total) using standard micro-costing methods described previously^14,15^. Resource costs are reported in four categories: 1) indirect costs (support staff/personnel, building costs, equipment, and office supplies, including education and outreach costs); events (physician visits, counselling visits, and Fibroscans; laboratory tests (HCV viral load modeled using the centralized laboratory pricing at the Synevo laboratory, HCV genotyping, HIV and HBV testing, pregnancy testing if applicable, PT/INR, blood counts, chemistries, and imaging); and medications (HCV, HBV, and HIV). Costs incurred solely for research purposes (e.g. obtaining informed consent, data entry, etc.) were not included.

We determined variable patient resource utilization from a de-identified dataset compiled from study participant case reporting forms and estimated resource utilization from average clinic site capacity and total annual visits. We then multiplied the quantity of each resource used by each patient by the associated unit cost to determine a total cost per patient. We evaluated the average resource utilization per patient by cost category and the four HCV treatment outcomes defined for the cost analysis and estimated 95% confidence intervals by outcome category. Cost estimates utilized data from a subset of all patients enrolled in the study; the subset included sequentially enrolled patients during the first six months of the enrollment period.

Study data were managed with HepatiC® (ABL SA, Luxembourg). The patient resource utilization matrix for this study was generated using SAS software, Version 9.3 (SAS Institute Inc., Cary, NC, USA). Estimates per patient treated and per successful outcome were made using the HE^2^RO Healthcare Costs and Outcome Model (http://www.heroza.org/researchtools/the-healthcare-cost-and-outcomes-model-hcom) (HE^2^RO, Johannesburg, South Africa). Economic models were generated with Excel 2011.

### Ethical considerations

The study was reviewed by the Ukrainian Institute on Public Health Policy IRB #1, Institutional Review Board (00007612) and the University of the Witwatersrand Human Research Ethics Committee (M17078). The Boston University Institutional Review Board approved analysis of a de-identified analytic dataset (H-37820). All participants provided written informed consent. The study is registered at ClinicalTrials.gov (NCT04038320).

## Results

### Enrolment and patient characteristics

We consented 943 anti-HCV positive participants and enrolled those eligible at the Gromashevsky and Kyiv Hospital between 26 March and 02 November 2018. As shown in Figure 2, 43 participants were excluded after consent, leaving 900 eligible for treatment. Of these, 868 were initiated on treatment, and 860 were assessed for SVR at 12 weeks after treatment. Of those initiated on treatment, 44.1% (383) were HCV mono-infected, 55.5% (482) HCV/HIV infected, 0.3% (3) HCV/HBV infected. Most HCV mono-infected participants were recruited at the Kyiv City Hospital, while most HCV/HIV co-infected participants were from Gromashevsky. Out of the 868 who started treatment, 87% (755) were PWID, of whom 60.7% (434) were on MAT; most PWID were seen at Gromashevsky. The majority of participants, 91.8% (797), were not cirrhotic (Table 1). The mean APRI Score was 0.5 (0.3-0.8).

**Table 1:** Patient characteristics by treatment site

| Characteristic | Gromashevsky<br>n=434 | Kyiv City Clinic<br>n=434 | Total<br>N=868 |
| --- | --- | --- | --- |
| Age (years) (Median, interquartile range) | 39 (35-43.7) | 39 (35-45.8) | 39(35-44.7) |
| Sex (%): Female | 142 (32.7) | 153 (35.3) | 295 (34.0) |
| Education beyond primary (%) | 285 (65.7) | 288 (65.4) | 573 (66.0) |
| Risk groups (%) |  |  |  |
| Non PWID | 77 (17.7) | 35 (8.3) | 113 (13) |
| PWID | 357 (82.3) | 398 (91.7) | 755 (87) |
| PWID on MAT | 243 (98.4) | 191 (91.8) | 434 (95.4) |
| Marital status: never married (%) | 348 (80.2) | 351 (80.9) | 699 (80.5) |
| Median BMI (Kg/m<sup>2</sup>) (IQR) | 23.8 (21.5-26.3) | 24.0 (21.9-26.6) | 23.9 (21.6-26.5) |
| Cirrhosis % |  |  |  |
| Compensated cirrhosis | 41 (9.5) | 30 (6.9) | 71 (8.2) |
| No cirrhosis | 393 (90.6) | 404 (93.1) | 797 (91.8) |
| Infection status (%) |  |  |  |
| HCV mono-infected | 133 (30.7) | 250 (57.6) | 383 (44.1) |
| HCV/HIV | 301 (99) | 181 (98.4) | 482 (55.2) |
| HCV/HBV | 0 (0.0) | 3 (1.6) | 3 (0.3) |
| Genotype (subtype) |  |  |  |
| 1 | 295 (68.0) | 348 (80.2) | 643 (74.1) |
| 2 | 14 (3.2) | 6 (1.4) | 20 (2.3) |
| 3 | 115 (26.5) | 76 (17.5) | 191 (22.0) |
| 4 | 0 | 2 (0.5) | 2 (0.2) |
| Indeterminant | 10 (2.3) | 0 (0.0) | 10 (1.2) |
| Mixed | 0 | 2 (0.5) | 2(0.2) |
| HCV RNA, Log<sub>10</sub>IU/mL (median, IQR) | 6.9 (6.0-11.5) | 5.9 (5.5-6.2) | 6.1 (5.7-7.1) |
| CD4 T cell count | 515 (340-725) | 500 (374-675) | 506.5 (351-704) |
| Haemoglobin, g/dl (median IQR) | 14.7 (13.6-15.6) | 14.8 (13.5-15.9) | 13.5 (15.7) |
| Creatinine, mg/dL (median, IQR) | 71 (61-81) | 70 (61-80) | 70 (61-80) |
| ALT, U/L (median, IQR) | 51.9 (37-85.6) | 51.9 (37-85) | 53 (36-88) |
| Bilirubin, mg/dL (median, IQR) | 8.9 (6-13.4) | 10.1 (7.2-14) | 9.7 (6.6 -13.7) |
| Albumin, g/dL (median, IQR) | 47 (44.6-49.5) | 48.7 (46.6-51) | 47.9 (45.5-50.3) |
| INR (Median IQR) | 1 (0.9-1.0) | 1 (1-1.03) | 1 (1-1.03) |
| Platelets (x10<sup>9</sup>/L) (median (IQR)) | 211 (170-257) | 219 (186-255) | 215 (179 -255) |
| APRI score (median IQR) | 0.5 (0.3-0.9) | 0.5 (0.3-0.8) | 0.5 (0.3-0.8) |
| Fibroscan |  |  |  |
| F0-F1 | 2 (0.6) | 253 (71.5) | 255 (36.4) |
| F1-F2 | 34 (9.8) | 51 (14.4) | 328 (46.8) |
| F3 | 34 (9.8) | 36 (10.2) | 70 (10) |
| F4 | 0 (0.0) | 6 (1.7) | 9 (0.9) |

HCV = hepatitis C virus, PLHIV = people living with human immunodeficiency virus (HIV) infection, MSM = men who have sex with men, Partner + = sexual partner of HCV infected person, SW = sex worker, PWID = people who inject drugs, MAT= medication assisted treatment, HBV = hepatitis B virus, INR = International Normalized Ratio

**Figure 2:**
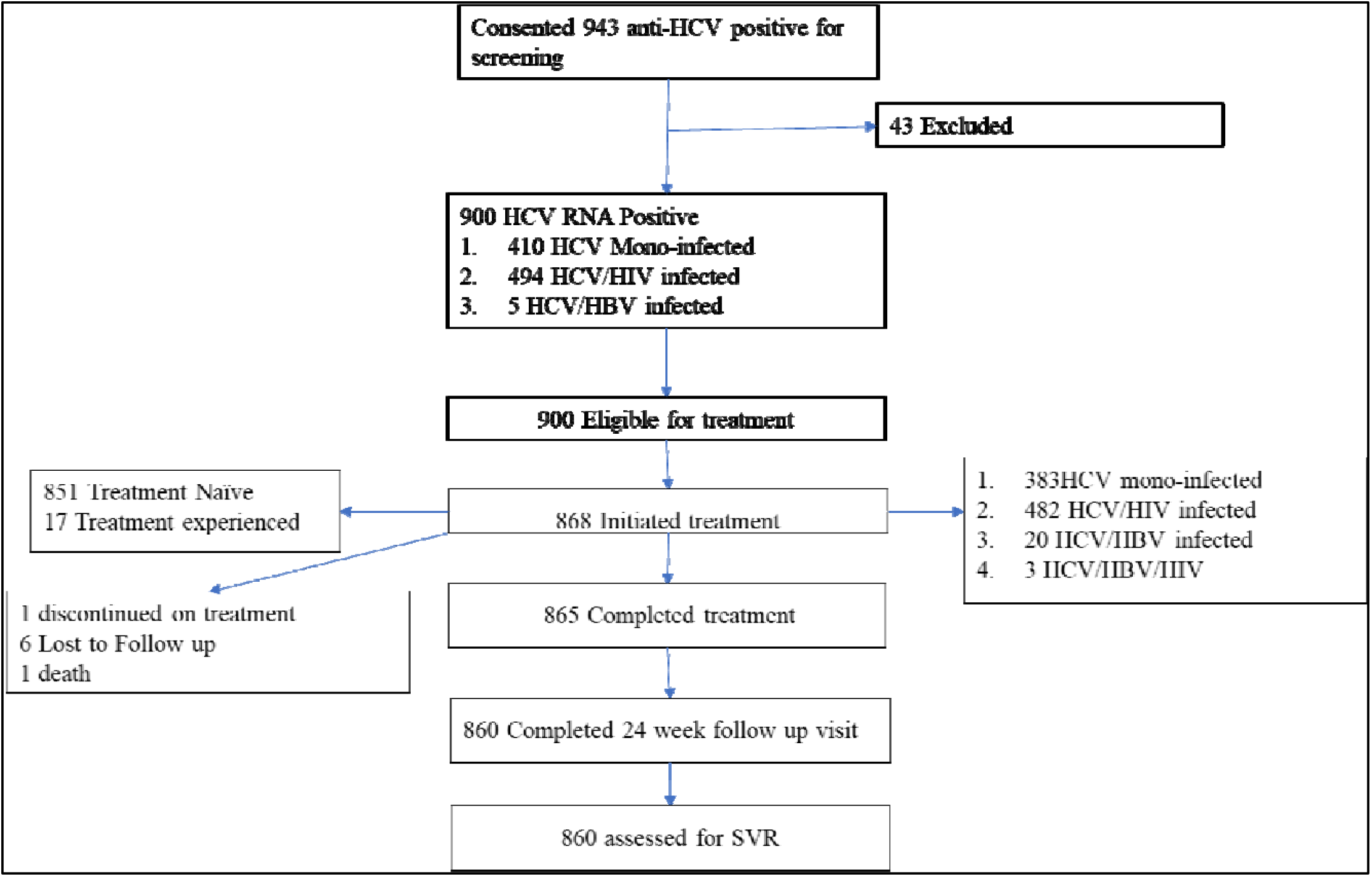
Enrolment of participants and outcomes

The most common HCV genotypes were 1 (74.1%) and 3 (22.0%) (Table 1). The median (interquartile range, IQR) HCV RNA was 6.1 (5.7-7.1) log_10_IU/m. Among those HIV infected (n=482), the median CD4 count was 507 cells/mm^3^ (351-704), and majority had HIV viral suppression (<200 copies per millilitre of blood) at entry. 20 had detectable viral load with a median (interquartile range) of 72 (52.5-267) copies per millilitre of blood; and 5 were unsuppressed (>200 copies per millilitre of blood). At 24 weeks, 12 weeks after the completion of HCV treatment, only two had unsuppressed viral loads.

### Treatment outcomes

Treatment outcomes by site are reported in Table 2. Of the 868 patients who initiated HCV treatment, 214 were treated with ribavirin. There were 80 adverse events reported, with 29, 20, 19 and 12 events at weeks 4, 8, 12 and 24 weeks, respectively. Of the 80 adverse events, 76.5% were mild and the rest moderate; 92.6% were either anaemia or hyperbilirubinemia, 3 were due to skin rash and itching, and 1 had lobar pneumonia. The majority 86.3% 69) were deemed study drug related; 89.9% (62/69 Ribavirin related events were managed by reduction of the dose and all SOF/LDV events were self-resolved.

**Table 2:** Outcomes at 24 weeks after study enrolment, by site

| Outcome (n, %) | Gromashevsky | Kyiv City Hospital # 5 | Total |
| --- | --- | --- | --- |
| Treatment success (SVR) | 427 (98.4%) | 404 (93.1%) | 831 (95.73%) |
| **Treatment failure**<br>(retained in care) | 0 (0%) | 29 (6.7%) | 29 (3.34) |
| Loss to follow up after<br>treatment initiation | 6 (1.4%) | 0 (0%) | 6 (0.69) |
| Premature<br>discontinuation | 1 (0.2%) | 0 (0%) | 1 (0.12) |
| Death | 0 (0%) | 1 (0.2) | 1 (0.12) |
| Total | 434 (100%) | 434 (100%) | 868 (100%) |

Following treatment initiation, 6 participants were lost to follow up (LTFU), 1 discontinued treatment prematurely and 1 died due to myocardial infarction between week 12 and week 24. Eight hundred and sixty were assessed for SVR at 24 weeks. By intention-to-treat analysis (ITT), which defined all non-successful outcomes as failures, 37 participants failed therapy, for an overall treatment success rate of 831/868 (95.7%). The SVR rates for participants with genotype 1 and 3 HCV were 95.5% and 96.3% respectively (Table 3). SVR rates differed significantly by site and by HIV coinfection status. The majority of treatment failures happened among the HIV-co-infected patients. All loss to follow up and the treatment discontinuation were from Gromashevsky clinic; the treatment failures were from Kyiv City Hospital (n=29 (Table 2). Of the 755 PWID assessed at 24 weeks, 95.2%. achieved SVR.

**Table 3:**
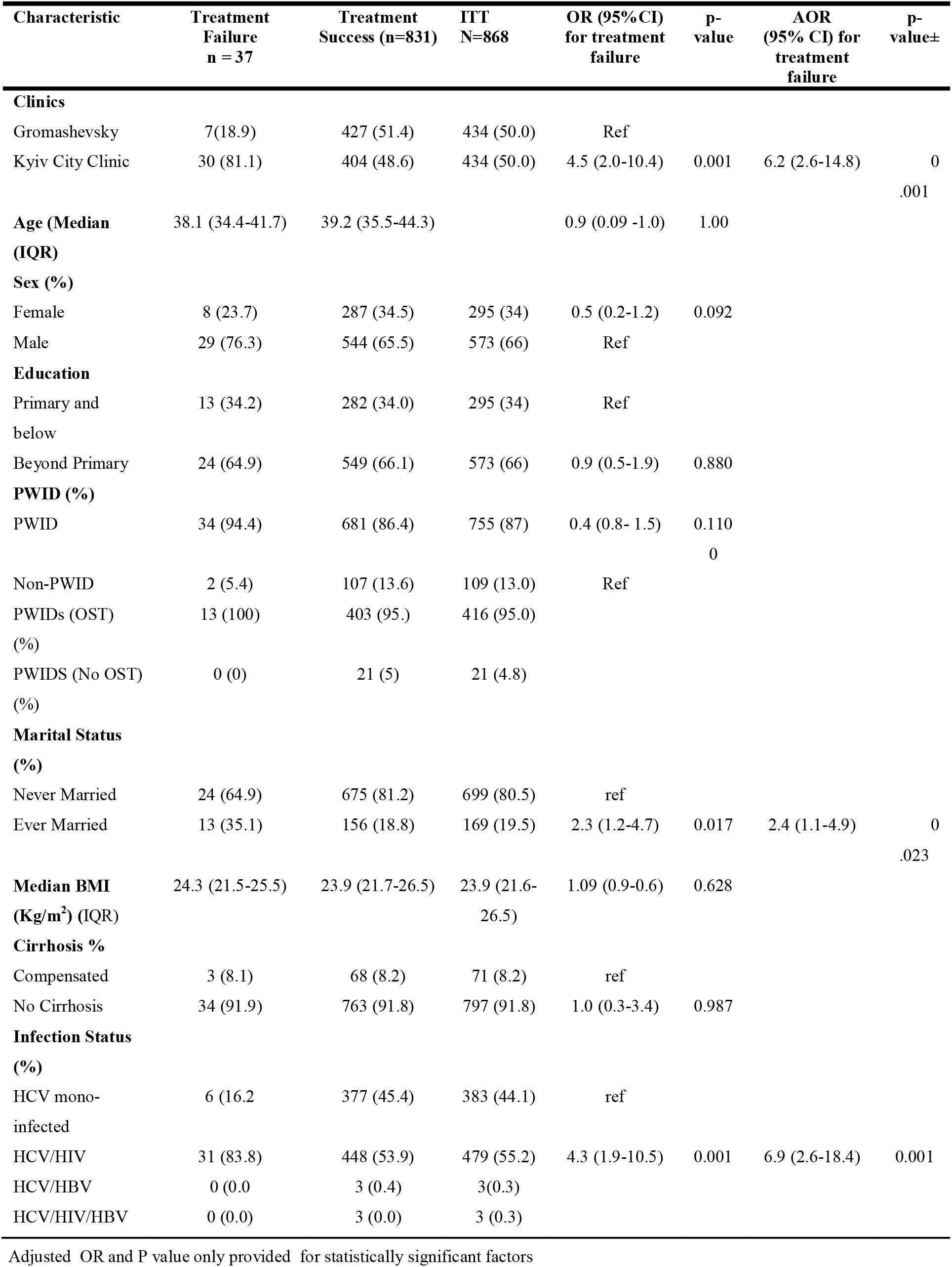
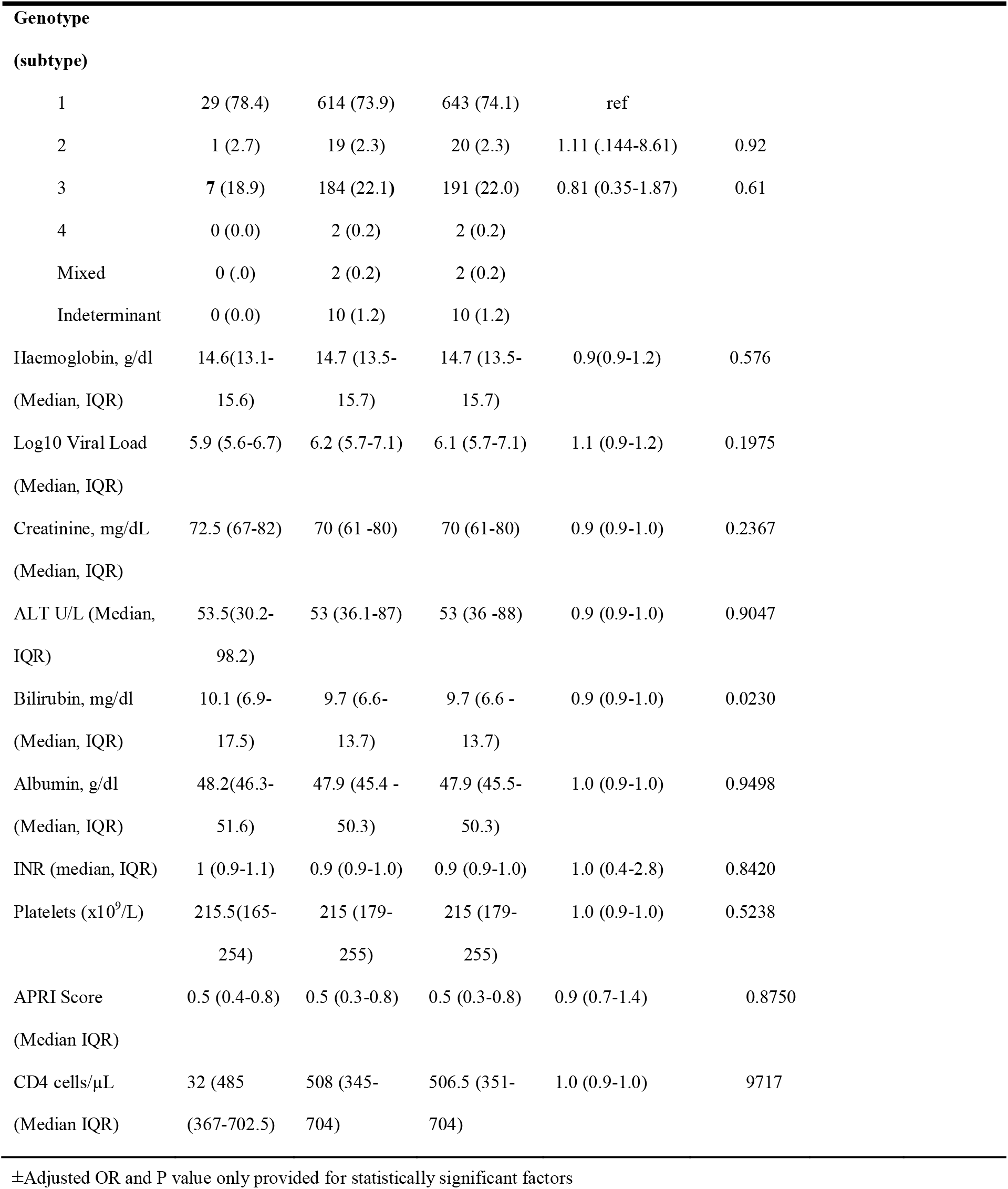
Factors influencing treatment response (intention to treat, N=868)

### Predictors of SVR

Characteristics of participants are compared between the SVR group and the non-SVR group by ITT population in Table 3. There were significant differences in the odds of achieving a successful treatment response by treatment site, marriage status, and HIV co-infection status in both univariate and multivariate analysis.

### Costs

Resource utilization for the first 522 participants enrolled at the Kyiv and Gromashevsky study sites and locally collected unit costs are presented in Table 4, and a detailed breakdown by cost component and outcome is provided in Table 5. We note that the estimates in Table 5 assume generic pricing for LDV/ SOF of $1.06 per tablet; brand pricing for this medication is estimated to cost $10.61/tablet. The results shown in Table 5 are a reasonable proxy for costs under “real world” conditions, with a cost per patient of $680.

**Table 4.** Mean resource utilization and mean HIV and HCV medication cost (USD) per participant during the 24-week study period

| Resource | Mean utilized per patient | Unit cost (2018 USD) |
| --- | --- | --- |
| *Resource quantities utilized and unit costs* |  |  |
| Physical exam | 2.0 | \$4.04 |
| Counseling visit | 6.0 | \$3.91 |
| Pregnancy test | 0.6 | \$1.00 |
| Fibroscan | 1.0 | \$45.23 |
| HCV RNA | 1.8 | \$33.71 |
| HCV genotype | 1.0 | \$25.31 |
| Liver tests (ALT, AST, albumin, bilirubin) | 4.1 | \$5.80 |
| CBC (Hb, Plt) | 3.7 | \$5.30 |
| Creatinine | 4.4 | \$1.40 |
| Urea | 1.5 | \$1.40 |
| INR/PT | 2.0 | \$2.70 |
| HIV screening antibody (OraQuick) | 1.0 | \$1.01 |
| CD4 count | 0.86 | \$10.00 |
| HIV RNA test | 1.0 | \$17.00 |
| HBV testing (Surface Ab, Core Ab, Surface Ag) | 1.0 | \$1.86 |
| *Medication costs* |  |  |
| HIV medication (mean/HIV+ participant per 24-week study period) | | \$298.96 |
| HCV medication using generic pricing (mean per 24-week study period) | | \$101.69 |
| HCV medication as procured in the study (mean per 24-week study period) | | \$912.29 |

HCV = hepatitis C virus, RNA = ribonucleic acid, ALT = alanine aminotransferase, AST = aspartate aminotransferase, CBC = complete blood count, Hb = hemoglobin, Plt = platelet count, INR = international normalized ratio, PT = prothrombin time, HIV = human immunodeficiency virus, HBV = hepatitis B virus, Ab = antibody, Ag = antigen

**Table 5.** Mean cost/patient by outcome in 2018 USD

| Outcome | N (%) | Indirect costs | Events | Labs | Drugs | Mean (95% CI) cost per outcome |
| --- | --- | --- | --- | --- | --- | --- |
| Success | 511 (98) | 174 | 77 | 171 | 257 | 679 (653-705) |
| Failure | 7 (1) | 174 | 65 | 182 | 206 | 627 (516-737) |
| Loss | 3 (1) | 141 | 77 | 145 | 243 | 606 (437-775) |
| Death | 1 (0) | 154 | 77 | 135 | 1,188* | 1,554 (N/A) |
| All outcomes | 522 | 173 | 77 | 171 | 258 | 680 (653-706) |

\*The patient who died was an HIV infected individual with Nevirapine (6 USD per pill) as a component of the antiretroviral regimen, incurring very high drug costs.

The study protocol included some resources that may not be regarded as essential for the intervention, such as a Fibroscan, and that many patients actually received more laboratory tests than were called for in the protocol. The cost of these excess resources is included in the estimate in Table 5. If the Fibroscan were removed and the quantities of liver tests, creatinine tests, and INR/PT were each cut in half, the average cost per patient would be $616.

## Discussion

In a demonstration project of an integrated, simplified protocol for treating HCV and HIV among key populations in Ukraine, of whom 87% (755) were PWIDS and 55.5% (482) were HCV/HIV co-infected, a high SVR rate of 95.7% was achieved amongst all who initiated treatment. Very high retention rates were observed especially in Kyiv City clinic. Treatment success varied by site, with Gromashevsky achieving 98.4 % SVR, and Kyiv city clinic 93.1%. This is unusual success in not only a high-risk population, but also for the efficacy of SOF/LDV +/-ribavirin particularly for genotype 3 infection. A prior US trial without ribavirin reported 89% SVR among genotype 3 patients without cirrhosis^16^, while a European study using ribavirin combination treatment reported and overall SVR of 94%^17^. While HIV co-infection was a risk for treatment failure, 93.6% of HIV/HCV co-infected individuals nonetheless achieved SVR. In addition only one HIV-coinfected patient was HIV viremic at 24 weeks. That may suggest the HIV/HCV co-infected who failed HCV treatment still adhered to HIV treatment.

Although PWID status was not associated with treatment failure, 91.9% (34) of those who failed were PWIDS. While adherence to HCV treatment was not measured, there was minimal loss to follow up; this may have been because of the Community Based Treatment model of care that incorporates a social worker into the clinical management team^18,19^ and also provides harm reduction services at a community level. Even among PWID, who typically have high rate of loss to follow up^20^, there were few losses, due perhaps to the Community Based Treatment harm reduction program in Ukraine^19^. Other studies have demonstrated high rates of success with HCV treatment co-located with substance use treatment services and amongst PWID receiving OST^21,22^.

Treatment with LDV/SOF and Ribavirin was generally well tolerated, with expected and manageable^23^ and non-requiring treatment discontinuation.

In our study, the estimated average cost of treatment per patient initiated on HCV therapy, including the costs for both HCV and HIV treatment, was $680. The small handful of patients who did not achieve SVR or did not return for the 24-week evaluation (LTFU) cost slightly less per patient, while the single patient who died cost substantially more, due largely to expensive HIV drugs The cost per patient treated reported here ($680/patient) is dependent on utilizing the lowest cost testing platform to confirm HCV viremia and treatment response (the status quo Synevo labs) and securing generic pricing for HCV medications. Other laboratory platforms will likely cause the cost/patient to go up or down depending on patient volume. Reducing the frequency of laboratory tests will also reduce costs. If generic drugs cannot be obtained, the cost of treatment will be substantially higher. As HCV treatment moves towards the use of pan-genotypic drugs, further reductions in the laboratory cost component are possible with elimination of genotype testing, though the cost of the medications will likely be higher than generic LDV/SOF. For program budgeting purposes, under the assumption of generic drug pricing but higher laboratory costs, an average cost of $750/patient is likely a reasonable estimate for this intervention. This does not include costs for scaling up or maintaining the treatment program, such as procurement, training, management, and oversight.

In conclusion, an integrated HCV/HIV treatment program in Ukraine achieved excellent outcomes with LDV/SOF+ribavirin HCV treatment in a patient population in which most individuals were co-infected with HIV and were PWID, and a substantial proportion had genotype 3 HCV infection. Loss to follow up was very low and cure rates very high. Adverse events due to RBV were minimal. The cost per patient, while relatively high for Ukraine, was modest by international standards. Going forward, a partnership among community-based organisations, private laboratories, and government that can secure generic medications and provide point of care testing may be an affordable strategy for making HCV treatment universally available in Ukraine.

## Data Availability

Data is available upon request

## Authors’ contributions

IS, SR, CC, KC, CVDH, TB, SA, conceived the study, FM, SA, YS, TB, ML, II, SM, MB conducted the study, CWK, CC, analyzed the data, CC, SR, AS and CWK drafted the manuscript and authors reviewed and approved the final manuscript.

